# Understanding the effectiveness of water, sanitation, and hygiene interventions: a counterfactual simulation approach to generalizing the outcomes of intervention trials

**DOI:** 10.1101/2022.11.15.22282349

**Authors:** Andrew F. Brouwer, Mondal H. Zahid, Marisa C. Eisenberg, Benjamin F. Arnold, Sania Ashraf, Jade Benjamin-Chung, John M. Colford, Ayse Ercumen, Stephen P. Luby, Amy J. Pickering, Mahbubur Rahman, Alicia N.M. Kraay, Joseph N.S. Eisenberg, Matthew C. Freeman

## Abstract

**Background:** While water, sanitation, and hygiene (WASH) interventions can reduce diarrheal disease, many large-scale trials have not found the expected health gains for young children in low-resource settings. Evidence-based guidance is needed to inform interventions and future studies.

**Objectives:** We aimed to estimate how sensitive the intervention effectiveness found in the WASH Benefits Bangladesh randomized controlled trial was to underlying WASH contextual and intervention factors (e.g.., baseline disease prevalence, compliance, community coverage, efficacy) and to generalize the results of the trial other contexts or scenarios.

**Methods:** We developed a disease transmission model to account for transmission across multiple environmental pathways, multiple interventions (water (W), sanitation (S), hygiene (H), nutrition (N)) applied individually and in combination, adherence to interventions, and the impact of individuals not enrolled in the study. Leveraging a set of mechanistic parameter combinations fit to the WASH Benefits Bangladesh trial (n=17,187) using a Bayesian sampling approach, we simulated trial outcomes under counterfactual scenarios to estimate how changes in intervention completeness, coverage, compliance, and efficacy, as well as preexisting WASH conditions and baseline disease burden, impacted intervention effectiveness.

**Results:** Increasing community coverage had the greatest impact on intervention effectiveness (e.g., median increases in effectiveness of 34.0 and 45.5% points in the WSH and WSHN intervention arms when increasing coverage to 20%). The effect of community coverage on effectiveness depended on how much transmission was along pathways not modified by the interventions. Intervention effectiveness was reduced by lower levels of preexisting WASH conditions or increased baseline disease burden. Individual interventions had complementary but not synergistic effects when combined.

**Discussion:** To realize the expected health gains, future WASH interventions must address community coverage and transmission along pathways not traditionally covered by WASH. The effectiveness of individual-level WASH improvements will be blunted the further the community is from the high community coverage needed to achieve herd protection.

## Introduction

Enteric diseases, primarily spread through contact with fecally contaminated environments (e.g., water surfaces, food), are one of the leading causes of morbidity and mortality in young children.^1^ An estimated nearly 500,000 children under five years of age die from diarrheal disease globally each year^2,3^, and it is hypothesized that repeated sub-clinical infections may lead to growth shortfalls.^4^ Much of this burden is in low- and middle-income countries (LMICs)^5^. Enteric pathogens are transmitted by myriad pathways, including fluids, fomites, food, flies, and fauna, as summarized in the classic “F-diagram”.^6^ Studying and preventing diarrheal disease is complicated because a diverse array of pathogens can cause similar symptoms,^7,8^ pathogens can exploit multiple transmission pathways, and asymptomatic infections can contribute to the community pathogen burden.^9^

Diarrheal disease is greatly reduced in communities with robust water, sanitation, and hygiene (WASH) infrastructure, with mutually reinforcing levels of community and individual protection. Household-leve WASH improvements can result in considerable reductions in diarrheal disease burden in LMICs, and many WASH interventions—such as improved latrines and handwashing with soap—have demonstrable efficacy to reduce fecal exposure.^10^ A recent meta-analysis by Wolf et al. of WASH intervention randomized controlled trials (RCTs) highlighted that WASH interventions reduce diarrhea in children in low-resource settings.^10^ However, there is substantial heterogeneity in the effect estimates across the studies, and many large-scale trials reported less-than-expected or null results.^11–17^ In particular, the results of the WASH Benefits (WASH-B) Bangladesh and Kenya trials and the SHINE trial, all of which found no impacts of WASH on linear growth but some mixed effects on diarrhea, were particular subjects of substantial discussion in the literature.^18–22^ The sub-optimal performance of the interventions in these trials is likely due to a combination of multiple factors, including incomplete blocking of all transmission pathways (low intervenable fraction, also called completeness), inadequate community coverage of the intervention, or a lack of intervention compliance or efficacy.^19,23^ Additionally, a community’s preexisting WASH conditions and baseline disease burden can also impact the real-world intervention effectiveness.^19,23^ Assessing which factors are the largest barriers to diarrheal disease reduction will aid policy-makers, practitioners, and researchers in deciding how best to invest in WASH programs and design the next generation of programs and trials.^18–24^

RCTs are considered the gold-standard for estimating causal relationships, and they are rigorous assessments of a particular intervention within a particular context at a particular point in time. But, their findings do not necessarily generalize to other contexts or conditions—e.g., different populations, disease burdens, pathogens, transmission pathways, intervention fidelity and adherence—when there are effect modifiers that vary across field settings and intervention implementations.^20,23^ Mechanistic infectiou disease transmission models, unlike meta-analyses, have the potential to generalize findings by directl accounting for these location-specific contexts and conditions, exploring counterfactual questions throug simulation of alternate scenarios, and developing location-specific programmatic targets. This approach is used extensively in other contexts to assess public health interventions or counterfactual conditions.^25–27^ A mechanistic, counterfactual approach could lead to better-targeted public health WASH interventions, policy recommendations, and field trials.^28,29^

The aims of this work are to evaluate hypotheses about what led to the sub-optimal reductions in diarrhea among intervention households in an RCT (WASH-B Bangladesh) using a compartmental transmission model and to provide a framework to support planning of WASH interventions and context-specific WASH programming. We previously developed this model framework accounting for multiple environmental transmission pathways, shared environments, pre-existing WASH conditions, and adherence to multiple interventions and applied it to the empirical trial data.^30^ Our approach generates thousands of combinations of coverage, intervention efficacy, and transmission pathway strengths that could reasonably underlie the trial results. In this analysis, we leveraged those parameter combinations to simulate how intervention effectiveness would have been different under alternate scenarios. These counterfactual simulations provide evidence for policy recommendations, programmatic targets, and an evaluation framework for next-generation WASH interventions.^20,21,24^

## Methods

### Summary of approach

In prior work, we developed a compartmental transmission model framework to explain the outcomes of a RCT.^30^ Then, we explored how effectiveness of a single intervention depends on six WASH factors:^31^

- *Preexisting WASH conditions* We account for the fact that a fraction of the population already has WASH infrastructure comparable to that provided by the intervention trial.
- *Disease transmission potential* We account for the baseline disease prevalence through the basic reproduction number *R*_0_, a summary measure of the disease transmission potential. Note that, given the values of the other WASH factors, there is a one-to-one correspondence between the disease prevalence and the basic reproduction number. In this analysis, we vary the baseline disease prevalence rather than *R*_0_, as it what intervention trials measure.
- *Intervention compliance* We account for both intervention fidelity (whether the intervention was delivered) and adherence (whether participants used the intervention), defining compliance as the fraction of participants assigned to an intervention who are using it.
- *Intervenable fraction of transmission* Any individual intervention targets some, but often not all, of the transmission pathways that pathogens exploit. We account for how much transmission is along pathways that the intervention attenuates (even if imperfectly) and how much even a perfect intervention would not affect.
- *Intervention efficacy* Because interventions do not result in perfect reduction of transmission or shedding, we account for the actual reduction.
- *Community coverage fraction* Intervention trials typically only provide the intervention to a subset of the population. We account for the fraction of the population is enrolled in the trial.

In this analysis, we use a multi-intervention version of the model to investigate outcomes in the WASH-B Bangladesh randomized controlled trial specifically. In prior work, we demonstrated how to find mechanistic parameter sets that were consistent with individual-level diarrheal outcomes.^30^ Here, we use these mechanistic parameter sets to simulate what intervention effectiveness would have been in each of the WASH-B Bangladesh trial arms under each of six counterfactuals corresponding to the six WASH factors above, accounting for uncertainty in the parameters underlying the real data (original scenario). By simulating what the intervention effectiveness would have been in the trial under alternative circumstances, we evaluated the extent to which each factor may have contributed to the observed outcomes.

### Data

The WASH-B Bangladesh trial was a cluster-randomized trial of the efficacy of water, sanitation, hygiene, and nutrition interventions, alone and in combination, on diarrhea prevalence and linear growth.^14^ The investigators measured (child-guardian-reported, past-seven-day, all-cause) diarrheal prevalence in children at three time points approximately one year apart. Households in the study area are typically organized into compounds in which a patrilineal family shares a common space and resources, such as a water source and latrine. A total of 5551 compounds were enrolled, contingent on having a pregnant woman in her second trimester during the enrollment period. The study followed one or more target children born after baseline, as well as any other children in the compound who were under age 3 at baseline. These compounds were grouped into 720 clusters. Each cluster was assigned to one of seven arms testing combinations of four interventions: water chlorination (W), a double-pit, pour-flush improved latrine (S), handwashing with soap and water (H), and supplementary nutrition sachets (N). The control arm (C) consisted of 180 compounds, while 90 were assigned to each of the water (W), sanitation (S), handwashing (H), nutrition (N), combined water, sanitation, and handwashing (WSH), and all interventions (WSH-N) arms. Specific details on trial design, interventions, and results are published elsewhere^14,32^. We assessed whether any individual was using an intervention or a substantively equivalent preexisting WASH condition through four indicators defined and assessed by the investigators: detection of free chlorine in water (W), latrine with a functional water seal (S), handwashing station with soap and water (H), at least 50% of nutrition sachets consumed (N). The W and H interventions were intended for the households of the target children, but we were not able to determine whether other children in the compound were in that household or not. For this analysis, we assumed that non-target children were covered by the interventions; any misspecification will attenuate the estimated efficacy of the W and H interventions.^14^ We removed individuals with negative reported ages (n=2), missing reported diarrhea (n=2,745), or missing in any of the four use indicators (n=2,660), which left 17,187 individual observations (76% of the original sample) over the three surveys.

### Ethics

This secondary analysis of deidentified data was not regulated as human-subjects research.

### Model

Our compartmental transmission model, denoted SISE-RCT, is a susceptible-infectious-susceptible (SIS) model with transmission through environmental (E) compartments and simulated to steady state to approximate an RCT.^30^ The SISE-RCT model accounts for the six key mechanistic factors underlying WASH RCT outcomes described above. We previously developed the single-intervention model,^31^ and we include a description of it in the supplemental material for convenience.

As discussed in the Data section above, the WASH-B Bangladesh trial included 720 clusters of households each assigned to one of seven arms (control, W, S, H, N, WSH, WSHN). We extended the single-intervention SISE-RCT model to a multi-intervention model by accounting for transmission across three environmental pathways (water, fomites & hands, and all others combined), four interventions applied individually (W, S, H, N) and in combination (WSH, WSHN), and individual-level compliance with interventions or preexisting conditions. In brief, we modeled each of the 720 clusters with susceptible and infectious compartments for each of 2^4^=16 combinations of interventions/conditions depending on household adherence, i.e., in every cluster, we modeled the infection prevalence for each combination of having or not having each intervention or equivalent preexisting WASH condition. For example, for a cluster in the WSH arm, we estimated how many people were not using any interventions, how many were using W only, how many were using S only, etc., and what the infection prevalence was among each group given their collective interaction through the shared environments.

Specifically, for a given cluster, we denote the fraction of the population that is susceptible to infection and is using intervention(s) or preexisting WASH condition(s) *i* in {0,*W,S,H,N,WS*, …, *WSHN*}as *S* _i_, where 0 indicates the use of no intervention or preexisting WASH condition. Analogously, we denote the fraction of the population that is infected analogously by *I*_i_. The intervention and control arms are simulated separately. The populations with regular and attenuated exposure are modeled in every cluster simulation, accounting for the fraction of the population 1) enrolled in the study (the community coverage, *ω)*, 2) with preexisting WASH conditions (*ρ*_0_), and 3) complying with the intervention (*ρ)*. Note that *ρ*_0_ and *ρ* are vectors of length 16 that each sum to 1; i.e., everyone is categorized into one of the 16 exposure groups. For each cluster, the overall fraction of the population in each exposure group is given by the vector

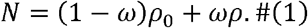

That is, the fraction of the population enrolled in the study (*ω)* follows the intervention compliance distribution of exposure groups (*ρ)*, and the fraction of the population not enrolled (1-*ω)* follows the preexisting WASH condition distribution of exposure groups (*ρ*_0_). In the control arm, *ρ = ρ*_0_.

The environment is partitioned into the three environmental pathways: water (*E*_*w*_), fomites & hands (*E*_*f*_), and all other pathways (*E*_*0*_). We assume that chlorination reduces transmission along the water pathway, sanitation reduces shedding into the water pathway, handwashing reduces transmission along the fomite pathway, and nutrition reduces susceptibility (transmission) for all three pathways. For each of the three pathways *j*, an environmental compartment *E*_*j*_ is characterized by the shedding into the environment (*α*_*j*_), the decay of pathogens in the environment (*ξ*_*j*_), and the transmission of pathogens from the environment to susceptible individuals (*β*_*j*_).The relative magnitude of shedding into *E*_*j*_ and relative transmission from *E*_*j*_ for the attenuated compared to the exposed populations are given by 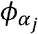 and 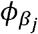, respectively. We also accounted for the possibility that the preexisting conditions were less efficacious than the RCT intervention. Once infected, individuals clear the infection at rate *γ*.

The model diagram of the multi-intervention SISE-RCT is given in Figure 1, although only two of the sixteen different exposure populations are shown. The full equations are given below (Eqs 2). A transmission term β_*j*_ *E*_*j*_ denotes transmission from the environmental pathway *j* The transmission term β_*j*_ *E*_*j*_ is attenuated by 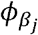 only for people in an attenuated exposure group (*S*_*i*_) using an intervention or preexisting condition that reduces transmission from pathway *j*, and contamination of that environmental pathway is attenuated by 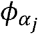 only for infectious people in an exposure group (*I*_*i*_) using an intervention or preexisting condition that reduces shedding into pathway In the following equations, the subscripts *w, f*, and *o* represent the water, fomite and other pathways. The subscripts *0, W, S, H, N*, and any combinations represent the exposure groups as defined above. Parameters *ω, ρ*, and *ρ*_0_ do not show up in these equations but are accounted for in the constraints, as discussed below. For brevity, we omit the 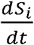 equations, each of which is given by 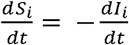 for the corresponding subpopulation.

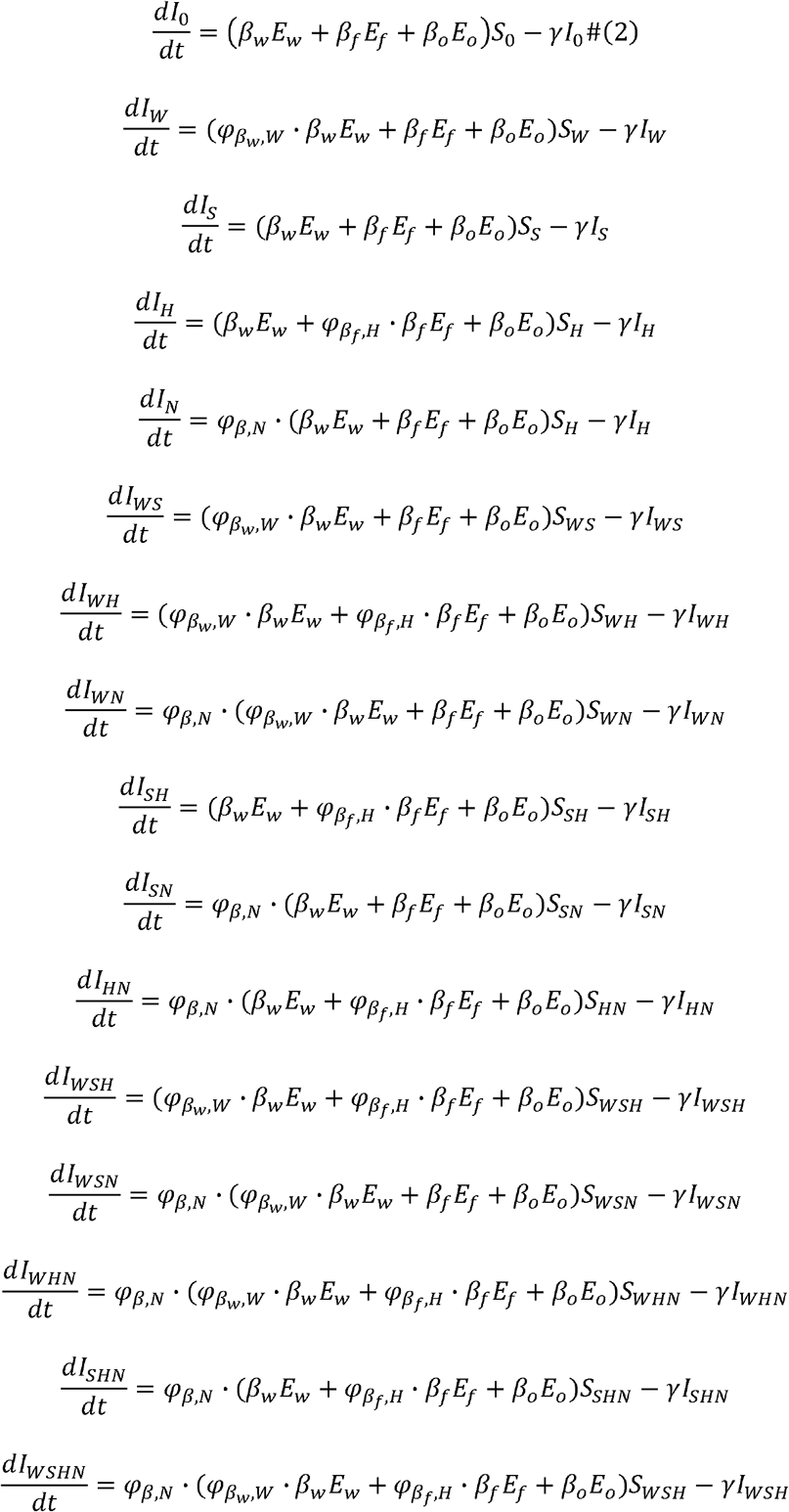

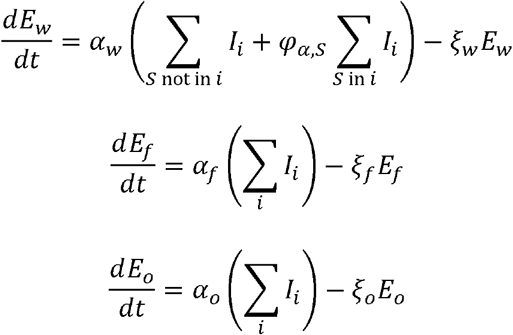

**Figure 1:**
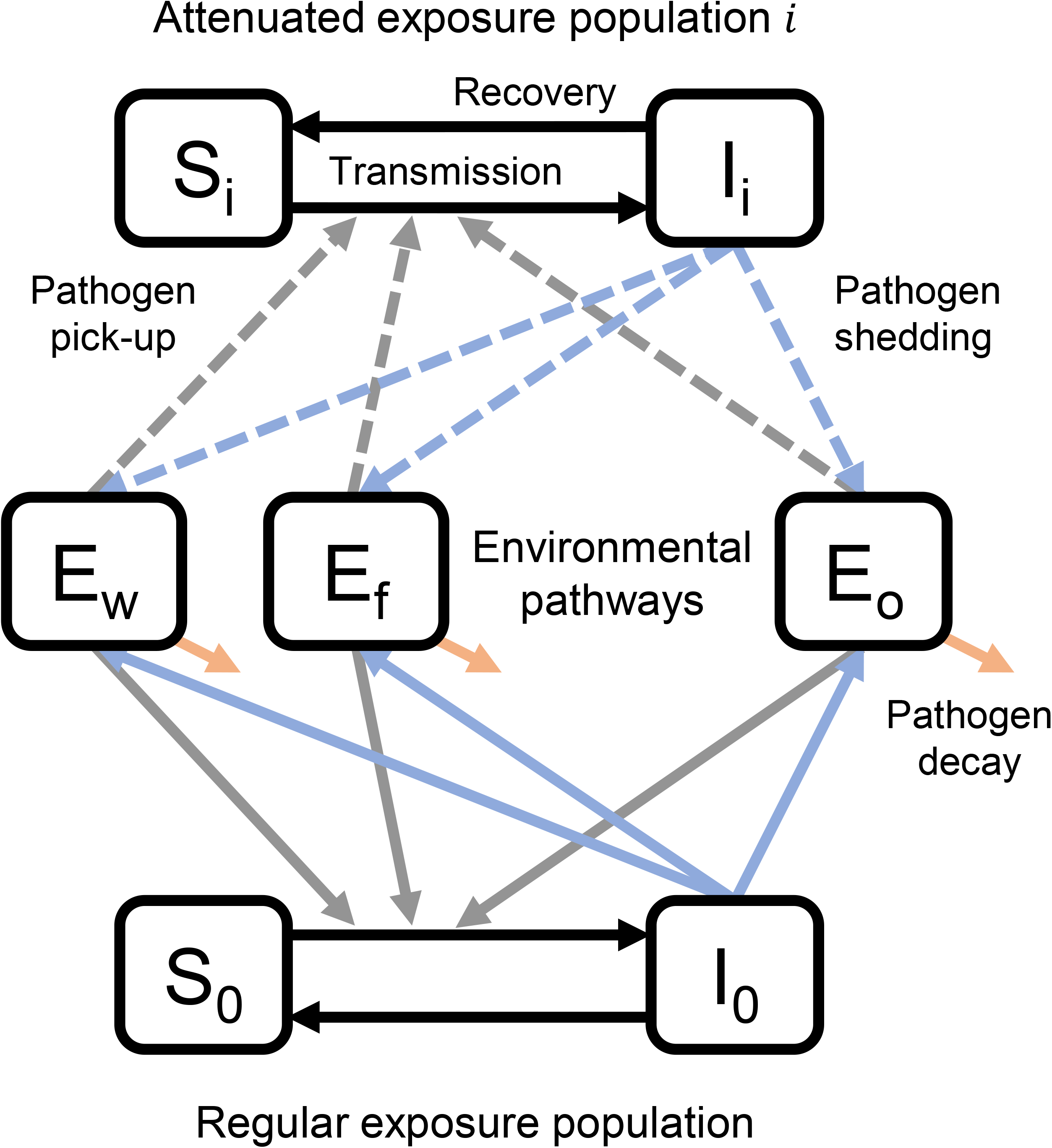
Single-intervention SISE-RCT model diagram with one attenuated exposure population and a regular exposure population interacting through shared environments. The SISE-RCT model is a compartmental susceptible-infectious-susceptible (SIS) model with transmission through environmental (E) compartments and simulated to steady state to approximate an RCT. The black lines denote infection and recovery, the blue lines denote shedding from infectious individuals into environmental compartments, the grey lines denote pick-up of pathogens from the environment by susceptible individuals, and the orange lines denote environmental pathogen decay. *S*_i_ and *I*_i_ denote susceptible and infectious fraction of the *i*th attenuated exposure population, and S_0_ and *I*_0_ denote susceptible and infectious fraction of the regular exposure population., *E*_*w*_ *E*_*f*_ and *E*_*o*_ denote environmental pathways for water, fomites & hands, and all other pathways. This figure was adapted from Figure 2 of Brouwer et al (2022).^30^

To find the steady state values (denoted by *) for the human compartments in the intervention arm, we set the above equations to 0 and simplify:

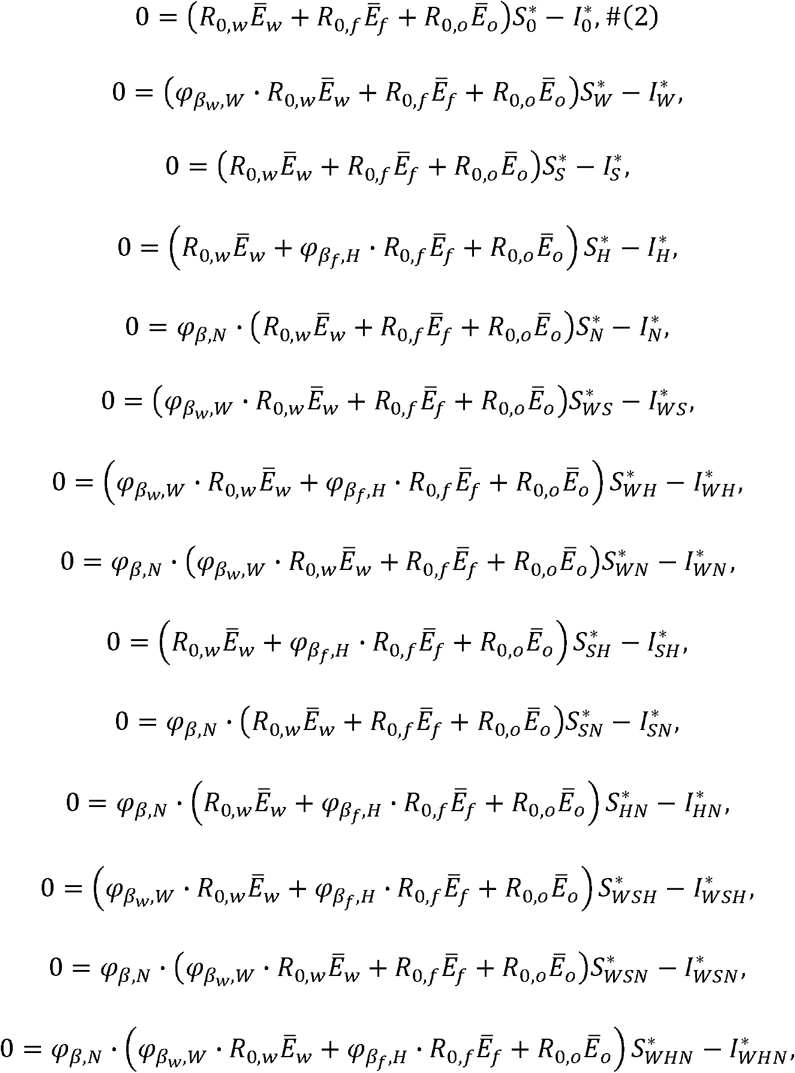

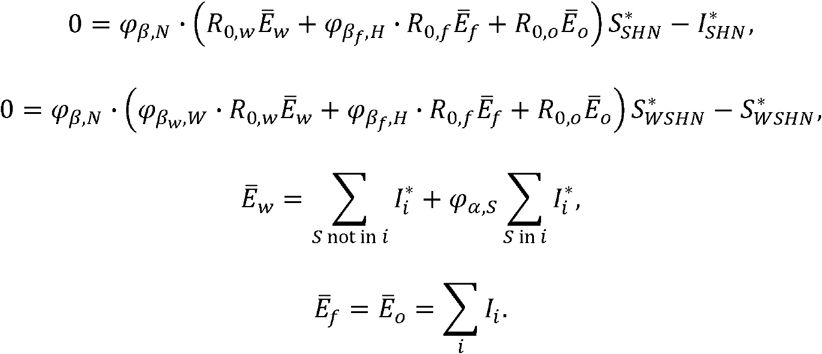

Here,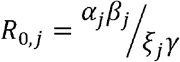 is the pathway-specific reproduction number for transmission through environment *E*_*i*_. The variables 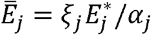 are conveniently scaled environmental steady states. For this model, the overall basic reproduction number is *R*_0_ = *R*_0, *W*_,+ *R*_0, *f*_ + *R*_0, *o*_, denoting the sum of the transmission potential through each pathway.

To solve for the steady states solutions for our 32 state variables (16 exposure groups times two susceptible/infection states), we solve the nonlinear system of equations (Eqs (3)) subject to the population constraint given in Eqs (1). The prevalence of disease in the population is denoted 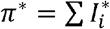. The prevalence in an intervention arm π^*^ is compared to the prevalence 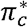 in the control arm, and the intervention effectiveness for that arm is defined as 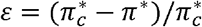, namely the fractional reduction in prevalence in the intervention arm relative to the control arm.

In summary, the model includes 18 parameters: i) the overall basic reproduction number *R*_0_, which defines the transmission potential in the control arm at baseline, ii) two parameters partitioning *R*_0_ into the strengths of the drinking water *R*_0,*W*_, fomite & hands *R*_0,*f*_, and all other transmission pathways *R*_0,*o*_, iii) eight relative reproduction numbers accounting for systematic differences in disease pressure over the trial time periods (baseline, midline, and endline) and across arms independently, iv) the community coverage *ω*, and v) efficacy parameters defining the effect of each intervention (four) or preexisting WASH condition (two) on the transmission pathways (the W intervention (chlorination) reduces transmission via the water pathway 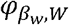 ; the S intervention (latrine with water seal) reduces shedding into the shared water environment with different efficacy for preexisting conditions 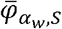 and the trial intervention 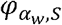; the H (handwashing with soap and water) reduces transmission via the fomite pathway with different efficacy for preexisting conditions 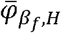 and trial intervention 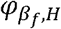; and the N intervention (nutrition supplementation) reduces susceptibility to all transmission *φ* _*β,N*_). In the Eqs (3), the basic reproduction number parameters are adjusted by the time and arm-specific and relative basic reproduction numbers corresponding to the cluster being modeled, and the intervention efficacy parameters 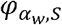 and 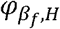 are replaced by 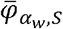 and 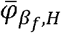 in clusters without the S and H interventions, respectively, and at baseline.

When solving for the steady state of these differential equations for a given cluster in a given time period we use the distribution of interventions and preexisting WASH conditions *ρ* recorded in the data for those participants and assume that participants not in the study have the same distribution of preexisting conditions as the control arm participants *ρ*_*0*_. We solved this system using the `nleqslv` package in R.

To fit the model to the trial data, we employed a hybrid sampling-importance resampling and estimation framework to obtain 50,000 parameter combinations that represented a good fit to the diarrheal outcomes of each participant using a Bernoulli statistical likelihood.^33^ We resampled, with replacement, from our initial 50,000 parameter combinations, based on their goodness of fit; 3,774 unique parameter combinations were included in the final sample, with varying frequency. These parameters sets are similar to but not exactly the same as in Brouwer et al,^30^ as they include a small code correction and use the computational improvement described previously.^31^ The fit to the data is given in Figure S1, and the distributions of parameters are given in Figures S2-5.

### WASH-B Bangladesh counterfactual analysis

We conducted two types of counterfactual analysis. First, we estimated the counterfactual intervention effectiveness in each arm across a range of each of the WASH factors starting from the scenario based on the median value of each parameter (which resulted in a fit close to the best-fit and was more representative of the parameter distributions than the specific best-fit parameter set). Because we are using the actual preexisting WASH conditions and intervention compliance recorded for each individual in the trial, there is not a well-defined way to continuously scale these two factors. So, we only compare the actual simulation to a “no preexisting conditions” and “full compliance” counterfactual, respectively.

Second, to account for the uncertainty in the parameters underlying the actual intervention outcomes, for each of the 50,000 parameter sets *k* identified by fitting the model to WASH-B Bangladesh, we defined the corresponding original scenario matching the WASH-B Bangladesh trial outcomes and the corresponding intervention effectiveness 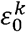. Because we are now accounting for uncertainty across these 50,000 parameters sets, it is not possible to succinctly capture changes as we continuously vary the factors. Thus, we considered six specific counterfactual scenarios, detailed in Table 2. Any parameter sets that eliminated disease in the control arm in a counterfactual simulation were censored from the results as they did not provide information on intervention effectiveness.

**Table 1:** Parameters of the SISE-RCT model. The SISE-RCT model is a compartmental susceptible-infectious-susceptible (SIS) model with transmission through environmental (E) compartments and simulated to steady state to approximate an RCT. The median value column denotes the median values for the multi-intervention model. Parameters *ρ*_O_ and *ρ* do not have median values because they are determined by the data.

| Parameter | Definition | Median value |
| --- | --- | --- |
| $\rho_0$ | Baseline WASH conditions (fraction of individuals in the community with intervention-level WASH infrastructure) | — |
| $\rho$ | Compliance (fraction of individuals in intervention arm using intervention) | — |
| $R_0$ | Transmission potential (basic reproduction number) | 1.10 |
| $\pi_c^*$ | Baseline disease prevalence (note: this technically a model output, but we treat it as a parameter because it has a 1-to-1 correspondence with $R_0$ given the other parameters) | 6.9% |
| $R_{0,w}/R_0$ | Fraction of transmission along the water pathway | |
| $R_{0,f}/R_0$ | Fraction of transmission along the fomite & hands pathway | |
| $R_{0,f}/R_0$ | Fraction of transmission on pathways not intervened on (i.e., 1 minus the intervenable fraction) | 0.48 |
| $1 - \varphi_\alpha$ | Intervention efficacy for reducing shedding (S=sanitation intervention) | 0.23 (S), |
| $1 - \varphi_\beta$ | Intervention efficacy for reducing transmission (W= water, H=hygiene, N=nutrition interventions) | 0.44 (W), 0.33 (H), 0.16 (N) |
| $\omega$ | Community coverage fraction (fraction of community included in the intervention trial) | 5.4% |

**Table 2:**
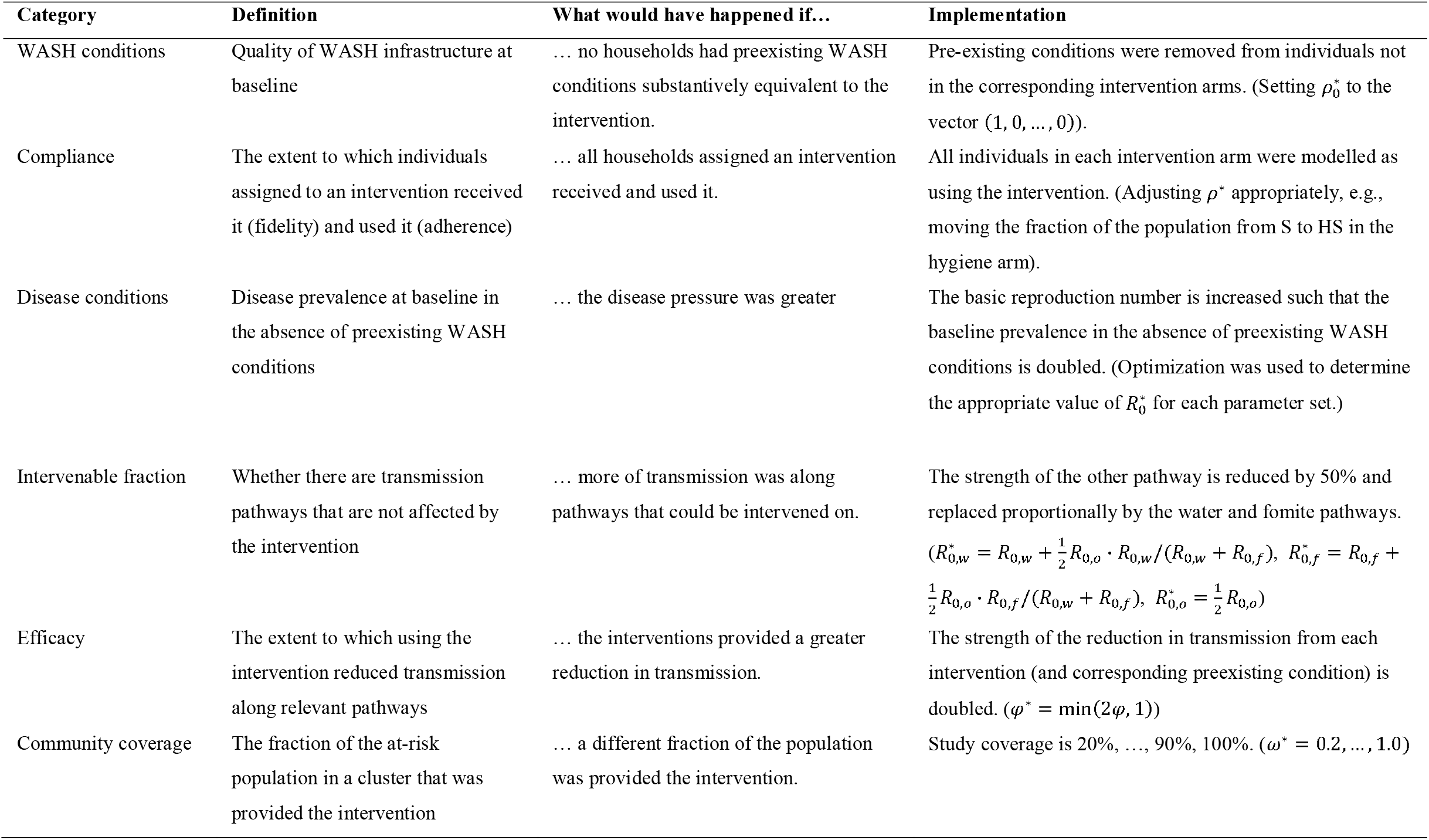
WASH-B Bangladesh counterfactual scenarios and implementations. WASH = water, sanitation, & hygiene. *indicates parameters in the counterfactual simulation.

The main outcome of a counterfactual simulation was the (absolute) change intervention effectiveness compared to the original scenario, namely 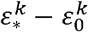, where 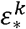 is the intervention effectiveness in the given counterfactual scenario for the th parameter set. We used absolute change rather than percentage change because absolute change, unlike percent change, is bounded (between -100 and 100 percentage points). The intervention is more effective (i.e., a greater reduction in diarrheal prevalence in the intervention arm compared to the control arm) in the counterfactual scenario than the original scenario when the change is positive. To assess whether the intervention factors modified the intervention effectiveness in the counterfactual scenarios, for a subset of counterfactuals, we assessed how the effect of the counterfactual depended on quantiles of the other parameter values.

The counterfactuals scenarios are not intended to be “plausible” for some specific, real-world changes. Indeed, changing the contextual factors of the preexisting WASH conditions and baseline disease prevalence is not possible. Instead, we can imagine these counterfactuals representing running the same trial in a different location to assess what the results would have been. The intervenable fraction is also not changeable for a given intervention (and underlying set of pathogens) but could be changed by adding additional intervention aspects to reduce transmission along other pathways. More broadly, we believe that investigating a broad range of counterfactual scenarios improves our understanding of the disease– intervention system, so that more effective interventions may be designed in the future.

## Results

### WASH-B Bangladesh counterfactual analysis from the median parameters

At the median parameter values (Figs S2-S5), intervention effectivenesses were 8.5% in the W arm, 40.8% in the S arm, 37.2% in the H arm, 32.1% in the WSH arm, 35.0% in N arm, and 34.6% in the WSHN arm, reflecting the results of the WASH-B Bangladesh trial. We estimated that removing all preexisting sanitation and hygiene infrastructure would have resulted in a modest reduction in baseline disease prevalence (by changing increasing the *R*_0_ parameter) nonlinearly decreased intervention intervention efficacy in all arms except for W, where the reduction was negligible (Figure 2a). Increasing baseline disease prevalence (by changing increasing th *R*_0_ 295 parameter) nonlinearly decreased intervention effectiveness in all arms, with decreasing reductions in effectiveness as baseline prevalence increased (Figure 2b). We estimated that there would be negligible-to-modest improvements in intervention effectiveness if there was full compliance (Figure 2c). Increasing the intervenable fraction (by reducing strength of the “other” pathway and proportionally increasing the strength of the water and fomite & hands pathways while keeping the overall *R*_0_ constant), increasingly improved the intervention the effectiveness (Figure 2d), with the WSH and WSHN interventions nearly achieving disease elimination if transmission were 100% intervenable. Increasing community coverage increased effectiveness approximately linearly, with each intervention achieving disease elimination at a different level of community coverage (Figure 2e; W: 75%, S: 90%, H: 75%, WSH: 35%, N: 55%, WSHN: 30%). Increasing intervention efficacy approximately linearly increased intervention effectiveness in the corresponding arms (Figure 2f-i). Increasing efficacy of the sanitation and hygiene interventions to 100% resulted in approximate disease elimination in the corresponding arms, but elimination would not have been achieved by increasing the efficacy of the W and N interventions.

**Figure 2:**
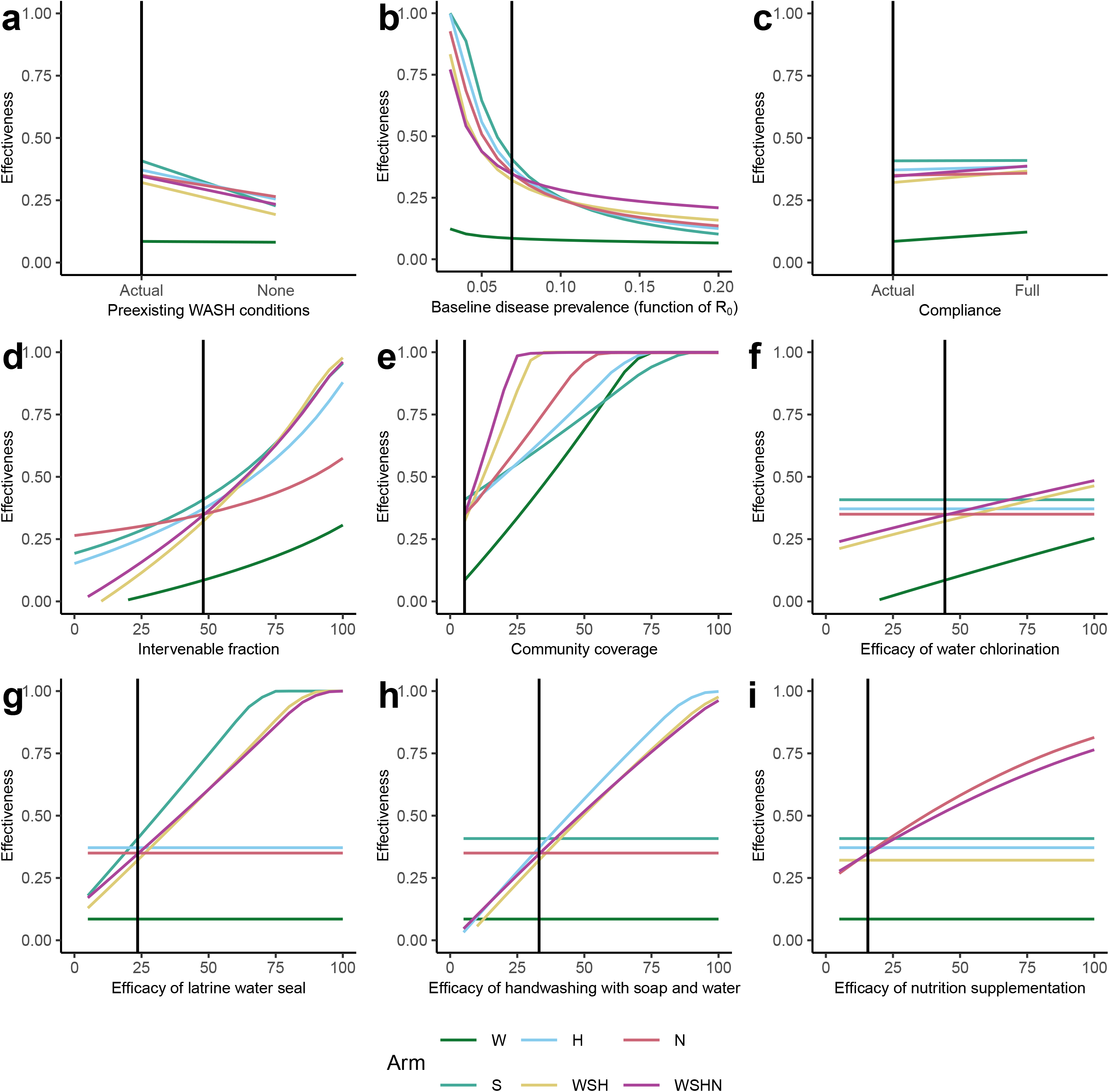
Intervention effectiveness as a function of WASH intervention factors. The SISE-RCT model is a compartmental susceptible-infectious-susceptible (SIS) model with transmission through environmental (E) compartments and simulated to steady state to approximate an RCT. The model was simulated at the median values of the model parameters when fit to the WASH Benefits Bangladesh trial and across ranges of counterfactual values of six contextual and intervention WASH factors. The six WASH factors are A) preexisting WASH conditions (fraction of individuals not enrolled in the intervention arm that are using preexisting WASH infrastructure), B) baseline disease prevalence (a function of the basic reproduction number *R*_O_), C) compliance (fraction of individuals enrolled in the intervention arm that are using the intervention), D) intervenable fraction of transmission (how much of the transmission could be prevented in a perfect intervention), E) the community coverage fraction (fraction of the population enrolled in the trial), and F-I) the intervention efficacy (fraction reduction in transmission or shedding when using the intervention) of each intervention. The underlying data are provided in Excel Table S1. WASH = water, sanitation, & hygiene.

### WASH-B Bangladesh counterfactual analysis accounting for parameter uncertainty

Because we do not know that the median values of the parameters in the scenario investigated in the first counterfactual analysis are accurate, we also considered the distribution of changes in intervention effectiveness when each counterfactual scenario (Table 2) was applied to the distribution of samples that fit the original data well. The median baseline disease prevalence in the original scenario was 7.1% (range 5.9–8.2%), decreasing to 5.7% (range 5.2–6.3%) for midline/endline (Figure S1). The median intervention effectivenesses were 8.2% in the W arm, 36.4% in the S arm, 33.1% in the H arm, 30.3% in the WSH arm, 34.0% in N arm, and 34.6% in the WSHN arm (Table 3). The percentage point change varied across arms in each counterfactual scenario. Figure 3 shows the distribution of percentage point change in intervention effectiveness over the 50,000 parameter sets for each arm and counterfactual scenario, and Table 3 gives the median values.

**Table 3.** Median intervention effectiveness and median percent point change in intervention effectiveness in each intervention arm for each counterfactual scenario compared to the original scenario. Intervention effectiveness (ε) in intervention effectiveness is 1 minus the relative risk of diarrhea in the intervention arm vs the control arm in each scenario, expressed as a percentage. The column Δε gives the median change in intervention effectiveness in percentage points (*not* the change in median intervention effectiveness); a negative number reflects a decrease in intervention effectiveness.

|  | W |  | S |  | H |  | WSH |  | N |  | WSHN |  |
| --- | --- | --- | --- | --- | --- | --- | --- | --- | --- | --- | --- | --- |
| | $\epsilon$ | $\Delta\epsilon$ | $\epsilon$ | $\Delta\epsilon$ | $\epsilon$ | $\Delta\epsilon$ | $\epsilon$ | $\Delta\epsilon$ | $\epsilon$ | $\Delta\epsilon$ | $\epsilon$ | $\Delta\epsilon$ |
| Original scenario | 8.1% | — | 36.3% | — | 33.0% | — | 30.2% | — | 33.9% | — | 34.5% | — |
| No WASH baseline conditions | 8.0% | -0.2% | 21.7% | -14.6% | 24.6% | -8.6% | 20.6% | -9.5% | 26.5% | -7.3% | 25.7% | -8.4% |
| Double baseline disease prevalence | 6.8% | -1.3% | 14.7% | -21.6% | 15.7% | -17.5% | 18.6% | -11.9% | 17.7% | -16.0% | 24.1% | -10.3% |
| Full compliance | 11.6% | +3.6% | 36.4% | +0.1% | 34.2% | +1.1% | 34.7% | +4.4% | 34.7% | +0.9% | 38.6% | +4.1% |
| Half of other pathway transmission can be<br>intervened on | 14.2% | +6.0% | 48.4% | +11.8% | 43.7% | +10.6% | 50.8% | +20.1% | 37.8% | +3.8% | 53.0% | +18.2% |
| Double efficacy of chlorination | 15.4% | +7.1% |  | — |  | — | 36.4% | +6.1% |  | — | 40.6% | +5.9% |
| Double efficacy of latrine water seal |  | — | 55.4% | +18.9% |  | — | 46.0% | +15.7% |  | — | 48.9% | +14.2% |
| Double efficacy of handwashing |  | — |  | — | 54.6% | +21.3% | 50.2% | +19.8% |  | — | 52.8% | +18.3% |
| Double efficacy of nutrition |  | — |  | — |  | — |  | — | 45.3% | +11.3% | 44.3% | +10.1% |
| Increase community coverage to 20% | 22.9% | +14.6% | 43.3% | +6.8% | 41.4% | +8.3% | 63.8% | +34.0% | 49.2% | +15.3% | 79.6% | +45.5% |

**Figure 3:**
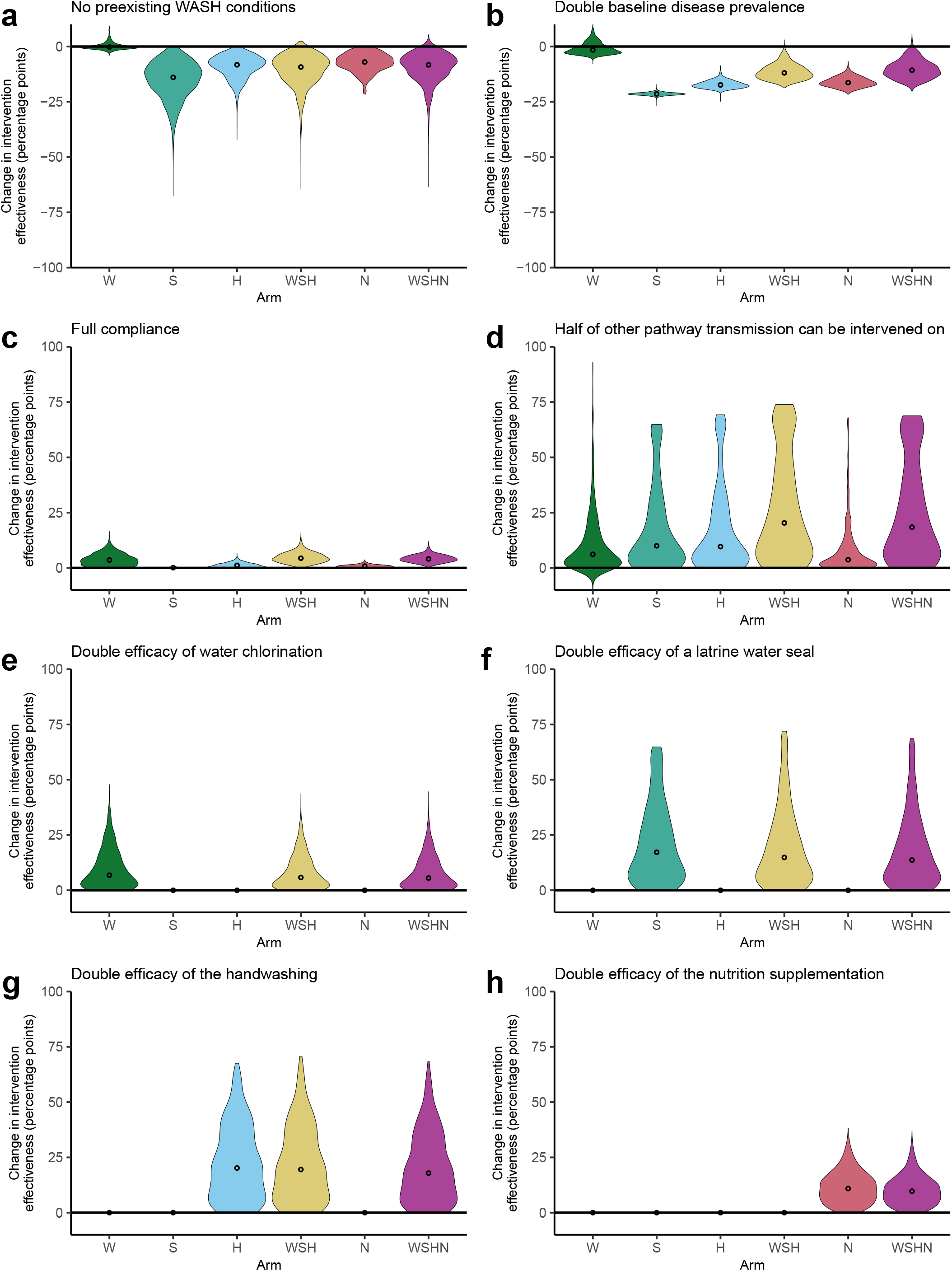
Percentage point change in intervention effectiveness compared to the original scenario in each counterfactual scenario. The SISE-RCT model is a compartmental susceptible-infectious-susceptible (SIS) model with transmission through environmental (E) compartments and simulated to steady state to approximate an RCT. Here, we applied it to data from the WASH-B Bangladesh trial, selecting 50,000 parameter sets consistent with the trial outcomes. We simulated each parameter set under each counterfactual scenario (Table 2). The violin plots give the distribution of values across the 50,000 simulations, with median points. The underlying data are provided in Excel Table S2A-H. W = water, S = sanitation, H = hygiene, N = nutrition.

#### Eliminate preexisting WASH Conditions

We found that implementing the interventions in a community with no handwashing stations with soap and water or latrines with water seals would have likely resulted in less effective interventions compared to the actual community’s higher baseline WASH conditions (e.g., 9.5 percentage points less in the WSH arm; Figure 3a). The W arm is the exception because it had lower effectiveness in the original scenario. The uncertainty in change in intervention effectiveness in each arm was largely driven by uncertainty in what the baseline disease prevalence would have been in the counterfactual scenario (median 8.9%, range 6.4–23.1%).

#### Double baseline disease prevalence

A higher transmission potential corresponding to a doubling of the baseline diarrheal disease prevalence (median 14.2% vs median 7.1%) would also have resulted in less-effective interventions compared to the true baseline diarrheal disease prevalence (e.g., 11.9 percentage points less in the WSH arm; Figure 3b). As above, the W arm is the exception because it had lower effectiveness in the original scenario.

#### Full compliance

The impact of increasing intervention adherence was negligible-to-modest (e.g., 4.4 percentage points more in the WSH arm; Figure 3c). Note that intervention compliance, as defined by the investigators, was already high.^14,34^

#### Half of the other pathway transmission can be intervened on

We found that intervention effectiveness could have been greater if more of the total disease transmission was via the water and fomite pathways rather than through pathways that were not intervened on (e.g., 20.1 percentage points more in the WSH arm; Figure 3d). There was potential for a substantial increase in intervention effectiveness, as indicated by the distribution of the individual simulation outcomes, but the median impact was modest, with a less than 25 percentage point increase in effectiveness in the multi-intervention arms and a less than 15 percentage point increase in the single-intervention arms (Table 3). The uncertainty in the potential impact was largely driven by uncertainty in how much of the disease transmission was through other pathways in the original scenario.

#### Double intervention efficacy

We assessed the impact of increasing efficacy—defined as increasing the reduction of transmission along the relevant pathway(s)—of the four interventions. We found that in each of these increased efficacy scenarios, substantial increases in intervention efficacy could have improved intervention effectiveness in the corresponding arms (Figure 3e–h), with median improvements between 5% and 20% points.

#### Increase community coverage

The median estimated community coverage in the trial was 5.4%, but this estimate was highly uncertain, ranging from nearly 0% to 20% (Figure S5). For our main coverage counterfactual, we increased the community coverage in each simulation to 20.0%, chosen as a substantial but not unreasonable increase in coverage.^35^ This counterfactual scenario was associated with the greatest median increase in intervention effectiveness (among all households now covered by the intervention) of any of the considered counterfactual scenarios (e.g., 34.0 and 45.5 percentage points more in the WSH and WSHN arms; Figure 4a). Following the results of the single-intervention model that highlighted that the effect of coverage depended on the other WASH factors, we plotted the intervention effectiveness distributions for this community coverage counterfactual by quintiles of the values of the other WASH factors. Effect modification is present if the effect of increased coverage depends on the quintile of the WASH factor. Note that when looking at quintiles of one factor, the values of the other factors may not be evenly distributed across the quintiles if values of the factors are correlated. We found that the increase intervention effectiveness with increased community coverage in the W, S, WSH, and WSHN intervention arms depended partly on the strength of transmission via the water pathway (Figure 4b). The increases in intervention effectiveness in these arms could only reach their full potential if the strength of the water pathway were high. A similar, but more modest effect was seen for the H arm and the strength of the fomite pathway (Figure 4c). The greatest overall effect modifier of the impact of increased coverage on intervention effectiveness is the strength of the other pathways (i.e., the intervenable fraction, Figure 4d). When the strength of other pathways was high, increasing coverage had less of an impact. Intervention efficacy also modified the impact of increased coverage but only in the intervention arms with those interventions (Figure 4e–h).

**Figure 4:**
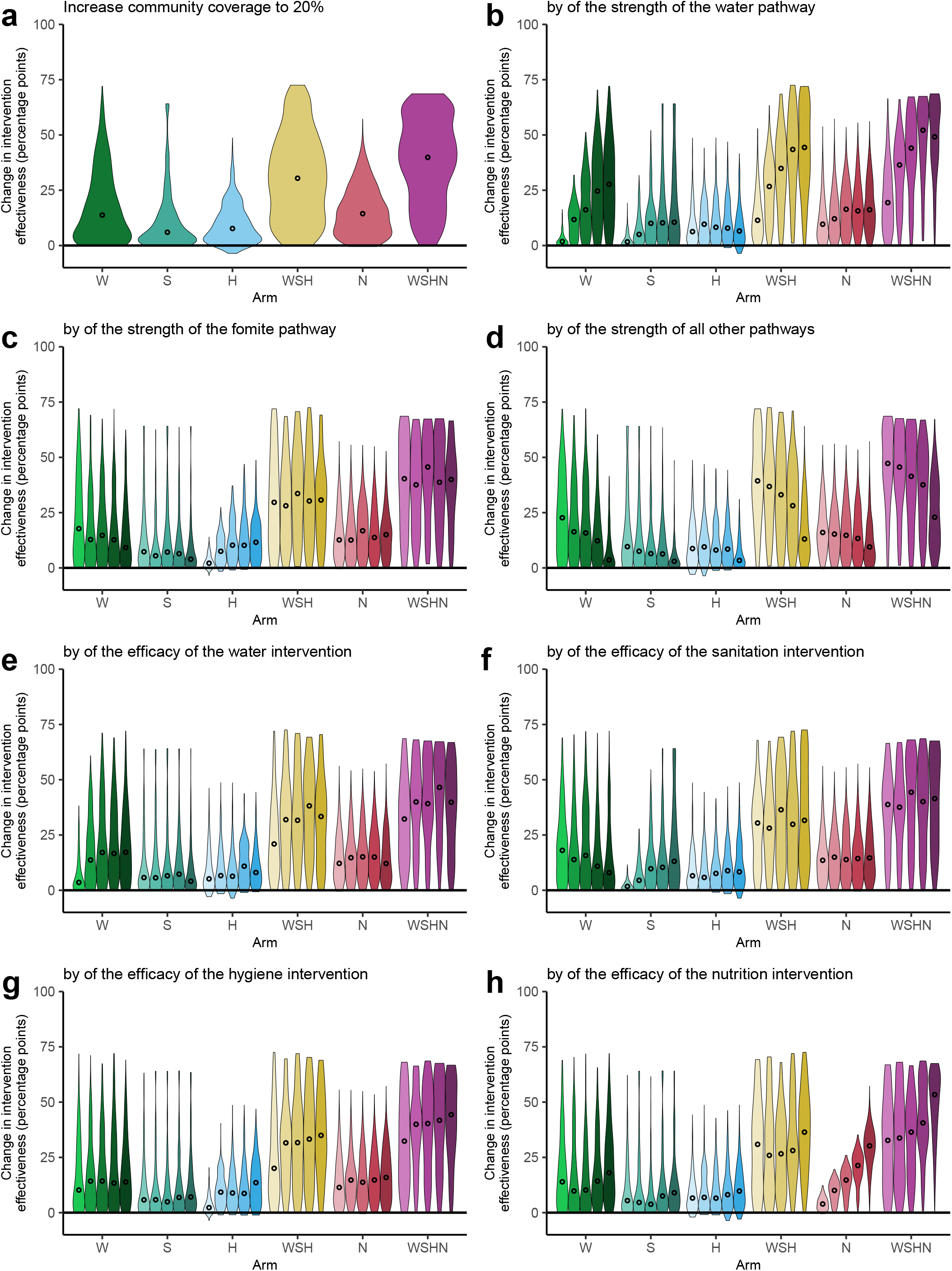
Percentage point change in intervention effectiveness compared to the original scenario in the 20% coverage counterfactual scenario (a) overall and (b) considering other parameters as potential effect modifiers (b-h). The SISE-RCT model is a compartmental susceptible-infectious-susceptible (SIS) model with transmission through environmental (E) compartments and simulated to steady state to approximate an RCT. Here, we applied it to data from the WASH-B Bangladesh trial, selecting 25,000 parameter sets consistent with the trial outcomes. We simulated each parameter set under the 20% coverage counterfactual scenario (Table 2). The violin plots give the distribution of values across the 25,000 simulations, with median points. In plots b-h, the five violin plots give the distributions of the intervention effectiveness across quintiles of the listed potential effect modifier. The underlying data are provided in Excel Table S3A-H. W = water, S = sanitation, H = hygiene, N = nutrition.

To further understand the joint impact of community coverage and the intervenable fraction (i.e., the strength of the other transmission pathway), we simulated the intervention effectiveness as a function of increased coverage for the highest and lowest quintiles of intervention completeness (Figure 5). The impact of increased coverage on intervention effectiveness depended on the intervenable fraction most strongly for the W, WSH, and WSHN arms, moderately for the S arm, and little for the H and N arms. For example, in the W arm, increasing community coverage to 50% resulted in a median increase of 80% points for samples with the highest intervenable fractions but only 23 percentage points for samples with the lowest intervenable fractions. In contrast, in the H arm, increasing community coverage to 50% resulted in a median increase of 31 percentage points for samples with the highest intervenable fractions compared only 22 percentage points for samples with the lowest intervenable fractions. (Note that the fact that intervenable fraction was relevant for the N arm at all is a result of the correlations between the intervenable fraction and the other parameters in the original parameter sets.)

**Figure 5:**
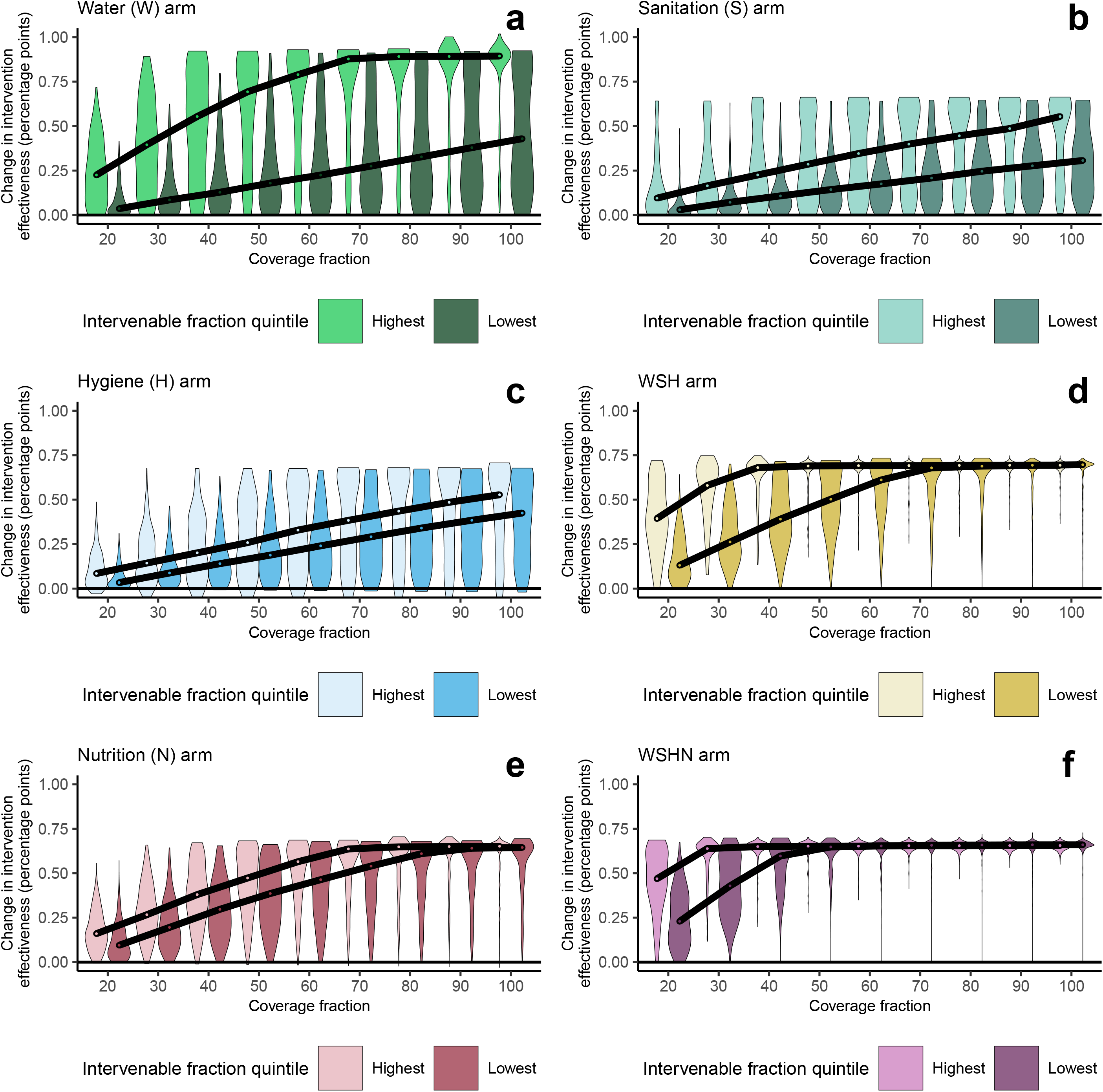
Percentage point change in intervention effectiveness compared to the original scenario in each arm for the lowest and highest quintiles of intervenable fraction. The SISE-RCT model is a compartmental susceptible-infectious-susceptible (SIS) model with transmission through environmental (E) compartments and simulated to steady state to approximate an RCT. Here, we applied it to data from the WASH-B Bangladesh trial, selecting 25,000 parameter sets consistent with the trial outcomes. We simulated each parameter set for coverage counterfactual scenarios ranging from 20% to 100% (Table 2). The violin plots give the distribution of values across the simulations, with median points and a line connecting the medians, for highest (dark) and lowest (light) quintiles of the intervenable fraction, i.e., the fraction of transmission that the interventions could directly act on. The underlying data are provided in Excel Table S4A-F. W = water, S = sanitation, H = hygiene, N = nutrition.

## Discussion

Our model-based analysis used counterfactual simulations to generalize the results of a WASH intervention trial and develop guidance for policymakers and researchers. Our first finding was that increasing community coverage led to the most substantial reduction in disease among people receiving interventions. Second, we found that intervention completeness (i.e., the fraction of disease transmission along pathways that were intervened on) was an important effect modifier of the impact of community coverage on intervention effectiveness, with the impact of increased community coverage enhanced when interventions covered a larger fraction of transmission. Third, our work suggests that interventions are likely to be more effective when disease burden is low. Finally, we found that multifaceted WASH interventions (WSH) added value over single component interventions (W, S, or H). Each of these findings suggest a path forward for policy and program recommendations for WASH investments and demonstrates how transmission models can be used to design the next generation of WASH interventions and set location-specific programmatic targets.

The importance of ensuring high community WASH coverage to address health outcomes has been highlighted in multiple context, including latrines,^29,36^ bed nets,^37^ and chemotherapy for helminths,^38^ among others. Further work is needed to improve our measures of indirect and direct intervention effects^29,39^ to better determine sanitation targets. Our findings support the call for systems-level WASH provisioning and improved universal access, underscoring the fundamental push to achieve the 2030 sustainable development targets.^40^

Our finding that the intervenable fraction (completeness) was an important effect modifier, emphasizes the need to better understand the sources of exposure not impacted by traditional WASH interventions. For example, contamination of food outside the home or from flies or exposure to feces from animals living near or inside the home may not be reduced by water quality or latrine interventions.^41–43^ Capturing and reducing transmission through additional targeted interventions would increase the fraction of transmission intervened on and thereby make increased community coverage even more effective.

Low diarrheal prevalence makes it more difficult to observe a statistically significant reduction in diarrhea.^14^ However, from a mechanistic perspective, we found that intervention effectiveness would have been lower had the background disease pressure in the community been higher (i.e., higher baseline disease prevalence) because individual-level interventions can be overwhelmed by higher disease pressure from the community, including those not covered by the intervention. This finding is supported by the outcomes of WASH-B Kenya trial, which had higher disease prevalence (27% in the control arm) and no significant intervention effects on diarrheal prevelance^15^ and is consistent with previous literature that has shown that non-pharmaceutical interventions are more effective for less transmissible pathogens or when the population has a higher degree of population immunity.^44^ This is not to say that individual improvements would have no effect but that the effects are blunted if disease pressure in the rest of the community were not also addressed.

Similarly, many have suggested that when preexisting WASH conditions are relatively high, interventions do not provide a substantial improvement in efficacy and thus health outcomes.^18,19,22,45^ However, from the transmission system perspective reflected by our results, if the preexisting WASH conditions (particularly among those *not* covered by the intervention) were poorer, the community disease pressure would be greater, and it would be more difficult to protect study participants from infection, even if the people covered by the intervention had a greater improvement in protection.

Because enteric pathogens can exploit multiple transmission pathways, many studies have tried to determine whether combined WASH interventions (WSH) are more effective than single interventions (W, S, or H).^32,46^ Whether or not there is an additional effect of combined interventions depends on whether the interventions are complementary, that is, whether they each block some of the transmission that the other interventions would not have blocked.^47^ This complementarity is an assumption in our transmission model framework (as each intervention affects different parts of the disease system), and because the model can fit the data, we find that complementarity is consistent with the observed trial results.^33^ Other modeling and empirical studies, support that WASH interventions can complement each other, or even potentially be synergistic.^48,49^ In this work, we found that the combined interventions could have a greater effect than the individual interventions, but that the effects were generally sub-additive, meaning that the effectiveness of the combined WSH intervention was less than the sum of its parts (Table 3). Combined interventions offer a substantially better chance of disease elimination, especially at higher coverage levels (Figure 5).

One challenge that WASH RCTs often face is achieving high compliance through both high fidelity (providing the interventions as planned) and high adherence of participants to the use of the intervention. In WASH-B Bangladesh, the intervention compliance, as defined by trial investigators, was high, generally above 90%.^14,34^ Accordingly, our full compliance counterfactual was limited in the impact it could detect.

The strength of our approach is underscored by the rich and high-quality data collected by the WASH-B Bangladesh trial (and other RCTs) and in our transmission model framework capturing relative disease prevalence. RCTs provide the gold standard of evidence about intervention effectiveness in a specific context, and our approach allows us to generalize RCT results to other contexts, providing a tool for powerful policy and programmatic guidance. The SISE-RCT model can be customized for local contexts and interventions and then used to support local decision-making (e.g., to determine whether to invest in community coverage vs intervention efficacy). Future work may also develop recommendations for achieving elimination while minimizing costs. One limitation of our study is the high uncertainty in many of the model parameters, especially the intervenable fraction, which propagates into the counterfactual scenarios. These uncertainties stem from potential trade-offs in the model, e.g., a low intervenable fraction and a low intervention efficacy may have similar effects. Fortunately, our framework has the potential to incorporate additional information about parameters like the intervenable fraction and efficacy through our Bayesian sampling-importance resampling approach, allowing us to tailor projections of intervention effectiveness to specific parameter regions based on additional information (e.g., chlorination efficacy above 75%). One limitation of the data was the inability to distinguish whether non-target children were members of the same household as the target child or not, which introduced misspecification into our classification of W and H exposures, likely attenuating the efficacy estimates for those interventions. Also, we accounted for changes in disease pressure between, but not within, survey periods; future work may more directly address seasonal changes in disease pressure and even pathway strength, as a function of precipitation, seasonal flooding, etc. Another limitation of this study is that our results do not directly address some aspects of the Sustainable Development Goal (SDG) Target 6.2.^40^ For example, the sanitation arm did not move households from no or basic sanitation to improved sanitation (as defined by the Joint Monitoring Programme). So, the “sanitation” intervention outcomes we estimated may not directly correspond to the policy-relevant changes required to meet SDG target. Likewise, the “water” intervention focused on water quality improvements (chlorination) but not water quantity. None of these issues are limitations of our modeling framework; rather, they are limitations of our specific application. Applying our methods across other trial datasets could address these limitations by allowing for modeling of other—and perhaps more policy-relevant—WASH exposure parameters.

Our work contributes to the robust discussion^18–24,50^ about the future directions of WASH research and programming, and our modeling approach is well-suited to reevaluating current evidence during the “pause for reflection” recommended by a consensus of WASH reserachers.^19^ This consensus group said that “the lesson perhaps lies in not seeking to attribute benefits to individual WASH factors but in that the public health dividends are paid when comprehensive services are in place.” Our work underscores this conclusion, not only by emphasizing the importance of coverage and completeness of interventions, but also in its rejection of the hypotheses that greater effectiveness might be found in areas with greater disease prevalence or lower preexisting WASH infrastructure. Indeed, our findings suggest that the effect of individual-level WASH improvements will be blunted the further the community is from achieving herd protection. Accordingly, this analysis provides further evidence supporting community-level interventions seeking to achieve herd protection through high community coverage.

## Data Availability

The WASH Benefits Bangladesh data is publicly available at https://osf.io/tprw2/. The data and code underlying the results of this paper are available at https://doi.org/10.5281/zenodo.10950560.

https://github.com/afbrouwer/WASH_RCT_transmission_modeling

https://doi.org/10.5281/zenodo.8057236

## Contributors

JNSE, MCE, MCF, and AFB conceived of the study. JNSE and MCF secured funding for the study. AFB, MCE, and JNSE developed the model. AFB wrote and implemented the software code, completed formal analysis and visualization, and curated the data and code. MHZ and ANMK validated the software code. AFB wrote the original draft with input from JNSE and MCF. BFA, SA, JBC, JMC, AE, SPL, AJP, and MR contributed equally by aiding in interpretation of the results and providing their expertise in the WASH Benefits trials. All authors reviewed and edited the manuscript. All authors had full access to all study data.

## Acknowledgements

This work was funded by the Bill & Melinda Gates Foundation (grant INV-005081) and the National Science Foundation (grant DMS-1853032). The original WASH-B Bangladesh trial was also funded by the Bill & Melinda Gates Foundation (grant OPPGD759). ANMK’s contributions were directly funded by the Bill & Melinda Gates Foundation and not as part of the foundation grant to the authors. ANMK is an employee of the Bill & Melinda Gates Foundation; however, this study does not necessarily represent the views of the Bill & Melinda Gates Foundation.

## Data sharing

The WASH Benefits Bangladesh data is publicly available at https://osf.io/tprw2/. The data and code underlying the results of this paper are available at https://doi.org/10.5281/zenodo.10950560. Data underlying each of the figures is given in the Excel spreadsheet supplement.

