## Supporting appendix for "Understanding the effectiveness of water, sanitation, and hygiene interventions: a counterfactual simulation approach to generalizing the outcomes of intervention trials"

**Single-intervention model**

Our compartmental transmission model, denoted SISE-RCT, is a susceptible-infectious-susceptible (SIS) model with transmission through environmental (E) compartments. To approximate the outcomes of a RCT, we solve for the model’s steady state in an endemic setting. The SISE-RCT model accounts for the six mechanistic WASH factors outlined above that underlie WASH RCT results. In the case of a single intervention, the population is partitioned into individuals with regular exposure (those not enrolled or included in the intervention and those not compliant), and those with exposure or shedding attenuated by the intervention (those compliant with the intervention or an equivalent preexisting WASH condition). Susceptible and infectious individuals with regular exposure are designated $S_{-}$ and $I_{-}$, and those with exposure or shedding attenuated by the intervention are designated$S_{+}$ and $I_{+}$. The intervention and control arms are simulated separately, and both the regular and attenuated exposure populations are modeled in both simulations, accounting for the fraction of population not enrolled in the study ($\omega)$, the fraction of the population with preexisting WASH conditions ($\rho_{0})$, and intervention compliance ($\rho)$. Individuals with regular exposure are either in the study but not compliant to the intervention ($\omega\left( 1-\rho\right)$) or are not in the study and do not have preexisting WASH conditions ($\left( 1-\omega\right)\left( 1-\rho_{0} \right)$). Individuals with attenuated exposure are either in the study and compliant to the intervention ($\omega\rho$) or are not in the study but have preexisting WASH conditions ($(1-\omega)\rho_{0}$). Hence, the population fractions of the attenuated and regular exposure populations are given by

$$\begin{aligned} N_{+}=\omega\rho+\left( 1-\omega\right)\rho_{0}, \#(1) \end{aligned}$$

$$\begin{aligned} N_{-}=\omega\left( 1-\rho\right)+\left( 1-\omega\right)\left( 1-\rho_{0} \right), \end{aligned}$$

respectively.

Once infected, individuals clear the infection at rate $\gamma$. An environmental compartment is characterized by the shedding into the environment $(\alpha)$, the decay of pathogens in the environment $(\xi)$ , and the transmission of pathogens from the environment to susceptible individuals $(\beta).$ For the single-intervention model, the environment is partitioned into the environmental pathway that is affected by the intervention $E_{1}$, either in terms of shedding into or transmission from the environment, and the environmental pathway that is not affected by the intervention $E_{2}$, with the same subscripts on $\alpha$, $\xi$, and $\beta$. For example, $E_{1}$ could be pathogens in water for an intervention that targets water, with $E_{2}$ representing all other potential transmission pathways (e.g., fomites, food, etc). The relative magnitude of shedding into $E_{1}$ and relative transmission from $E_{1}$ for the attenuated compared to the exposed populations are given by $\phi_{\alpha_{1}}$ and $\phi_{\beta_{1}}$, respectively.

The SISE-RCT parameters are given in Table 1, and a model diagram is given in Figure 1. The full equations are given below (Eqs 2). The two transmission terms $\beta_{1}E_{1}$ and $\beta_{2}E_{2}$ denote transmission from the environmental pathway attenuated by the intervention ($E_{1})$ and from the environmental pathway not attenuated by the intervention ($E_{2}$), respectively. The transmission term $\beta_{1}E_{1}$ is attenuated by $\phi_{\beta_{1}}$only for people in the attenuated exposure group ($S_{+})$, and contamination of that environmental pathway is attenuated by $\phi_{\alpha_{1}}$ only for infectious people of that same group ($I_{+})$. There is no attenuation of transmission to or shedding from the environmental pathway not affected by the intervention ($E_{2}$). Parameters $\omega,$ $\rho$, and $\rho_{0}$ do not show up in these equations but are accounted for in the constraints, as discussed below. For brevity, we omit the $\frac{dS}{dt}$ equations, each of which is given by $\frac{dS}{dt}= -\frac{dI}{dt}$ for the corresponding subpopulation.

$$\begin{aligned} \frac{dI_{+}}{dt}={(\phi}_{\beta_{1}}\beta_{1}E_{1}+\beta_{2}E_{2})S_{+}-\gamma I_{+},\#\#(2) \end{aligned},$$

$$\frac{dI_{-}}{dt}=\left( \beta_{1}E_{1}+\beta_{2}E_{2} \right)S_{-}-\gamma I_{-},$$

$$\frac{dE_{1}}{dt}={\alpha_{1}(\phi}_{\alpha_{1}}I_{+}+I_{-})-\xi_{1}E_{1},$$

$$\frac{dE_{2}}{dt}=\alpha_{2}\left( I_{+}+I_{-} \right)-\xi_{2}E_{2}.$$

To find the steady state values (denoted by *) for the human compartments in the intervention arm, we set the above equations equal to 0 and simplify out the environmental compartments,

$$\begin{aligned} 0={(\varphi}_{\beta_{1}}R_{0,1}(\varphi_{\alpha_{1}}I_{+}^{*}+I_{-}^{*})+R_{0,2}(I_{+}^{*}+I_{-}^{*}))S_{+}^{*}-I_{+}^{*},\#(3) \end{aligned}$$

$$0={(R}_{0,1}(\varphi_{\alpha_{1}}I_{+}^{*}+I_{-}^{*})+R_{0,2}(I_{+}^{*}+I_{-}^{*}))S_{-}^{*}-I_{-}^{*}.$$

Here, $R_{0,i}=\frac{\alpha_{i}\beta_{i}}{\xi_{i}\gamma}$ is the pathway-specific reproduction number for transmission through environment $E_{i}$. For this specific model, the overall basic reproduction number is $R_{0}=R_{0,1}+R_{0,2}$, denoting the sum of the transmission potential through the pathway affected by the intervention ($R_{0,1}$) and the pathway not affected by the intervention ($R_{0,2}$). The intervenable fraction (based on the strength of the transmission pathway targeted by the specific intervention) is $R_{0,1}/R_{0}$.

To get the steady states solutions for our four state variables ($S_{+}^{*}$, $I_{+}^{*}$, $S_{-}^{*}$, $I_{-}^{*}$), we solve the nonlinear system of equations (Eqs (3)) subject to the constraints $S_{+}^{*}+I_{+}^{*}=N_{+}$ and $S_{-}^{*}+I_{-}^{*}=N_{-}$, where $N_{+}$ and $N_{-}$ are given in Eqs (1). We solved this system using the nleqslv package in R. This approach is more computationally efficient than the differential equation simulation approach we used previously.^20^ We solve for the steady state in the control arm with the same parameters as the intervention arm except that $\rho=\rho_{0}$.

The prevalence of disease in the population is denoted $\pi^{*}=I_{+}^{*}+I_{-}^{*}$. The prevalence in the intervention arm $\pi^{*}$ is compared to the prevalence $\pi_{c}^{*}$in the control arm. Then, intervention effectiveness (the RCT outcome) is defined as $\varepsilon=(\pi_{c}^{*}-\pi^{*})/\pi_{c}^{*}$, namely the fractional reduction in prevalence in the intervention arm relative to the control arm.

**Multi-intervention model prevalence and parameter distributions**

A plot comparing the data in the WASH Benefits Bangladesh trial by wave (not combining midline and endline waves) to the posterior distributions of simulated prevalence using the SISE-RCT model is given below (Figure S1). The model is able to approximate the diarrheal prevalence in each arm at each time point, akin to how a linear regression line explains a set of points but does not intersect them all.

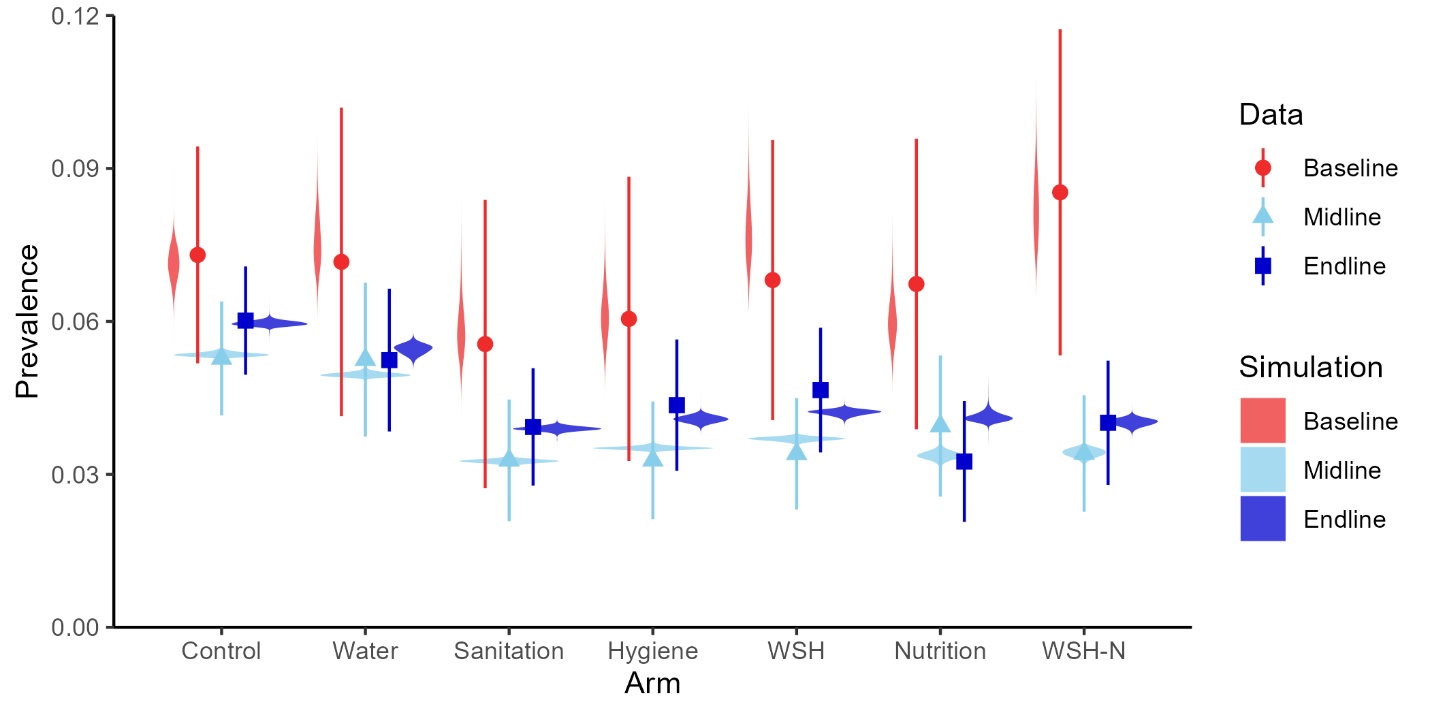

**Figure S1:** ***Prevalence****. Prevalence of self-reported diarrhea (7-day recall) in WASH Benefits Bangladesh comparing the baseline (red) to the combined midline (lightblue) and endline (darkblue) surveys, as well as posterior distributions of simulated prevalence (violin plots).*

We provide the posterior distributions for the basic reproduction number and pathway-specific reproduction numbers (Figure S2), the relative timepoint- and arm-specific basic reproduction numbers (Figure S3), the intervention and preexisting condition efficacy parameters (Figure S4), and the community coverage (Figure S5). These distributions are reproductions of those presented in Brouwer et al. (2022), correcting for a small code error and updating the computational framework.

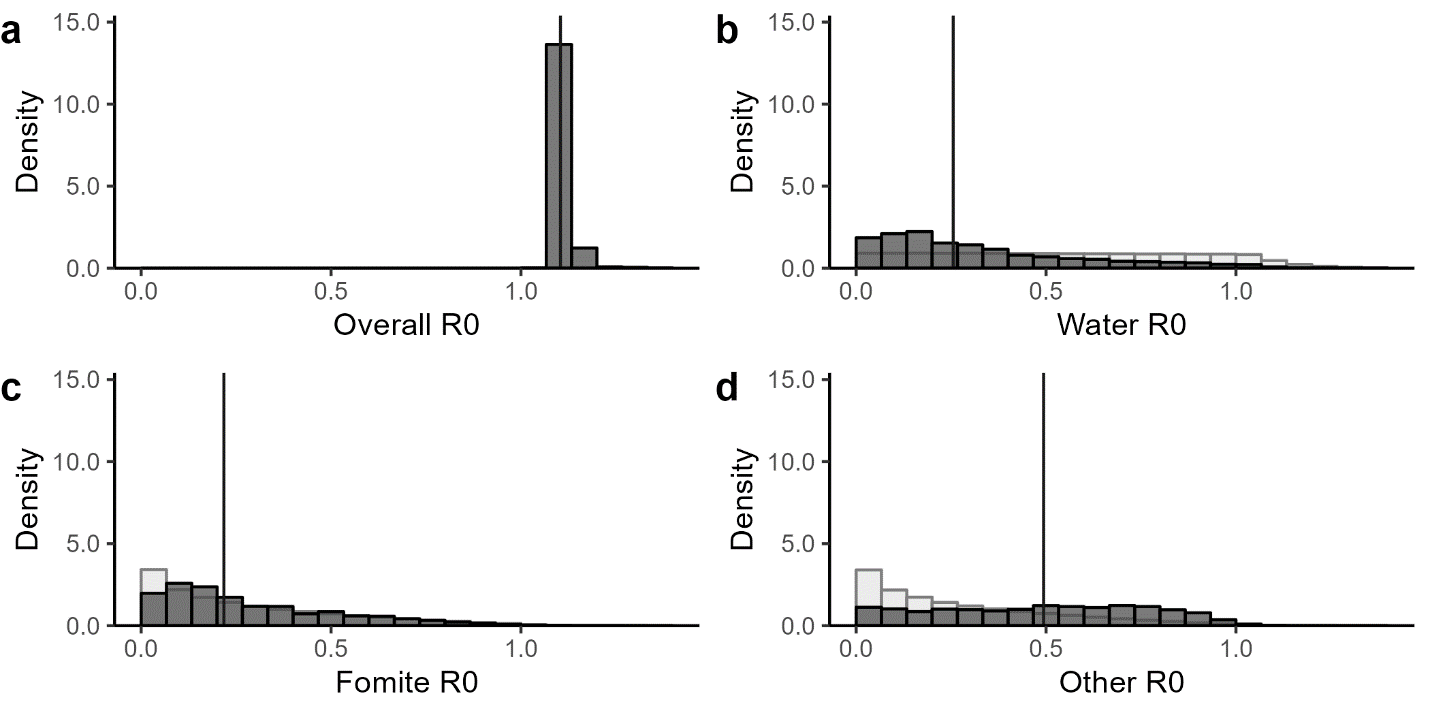

**Figure S2:** ***Reproduction numbers****. Posterior distributions (dark gray) of the (a) overall basic reproduction number R0, (b) the water pathway basic reproduction number, (c) the fomite pathway reproduction number, and (d) the other pathway reproduction number. Prior distributions (light gray) are given for the three sampled parameters; prior distributions are not uniform because they are products of underlying uniformly distributed parameters. Vertical lines give median values.*

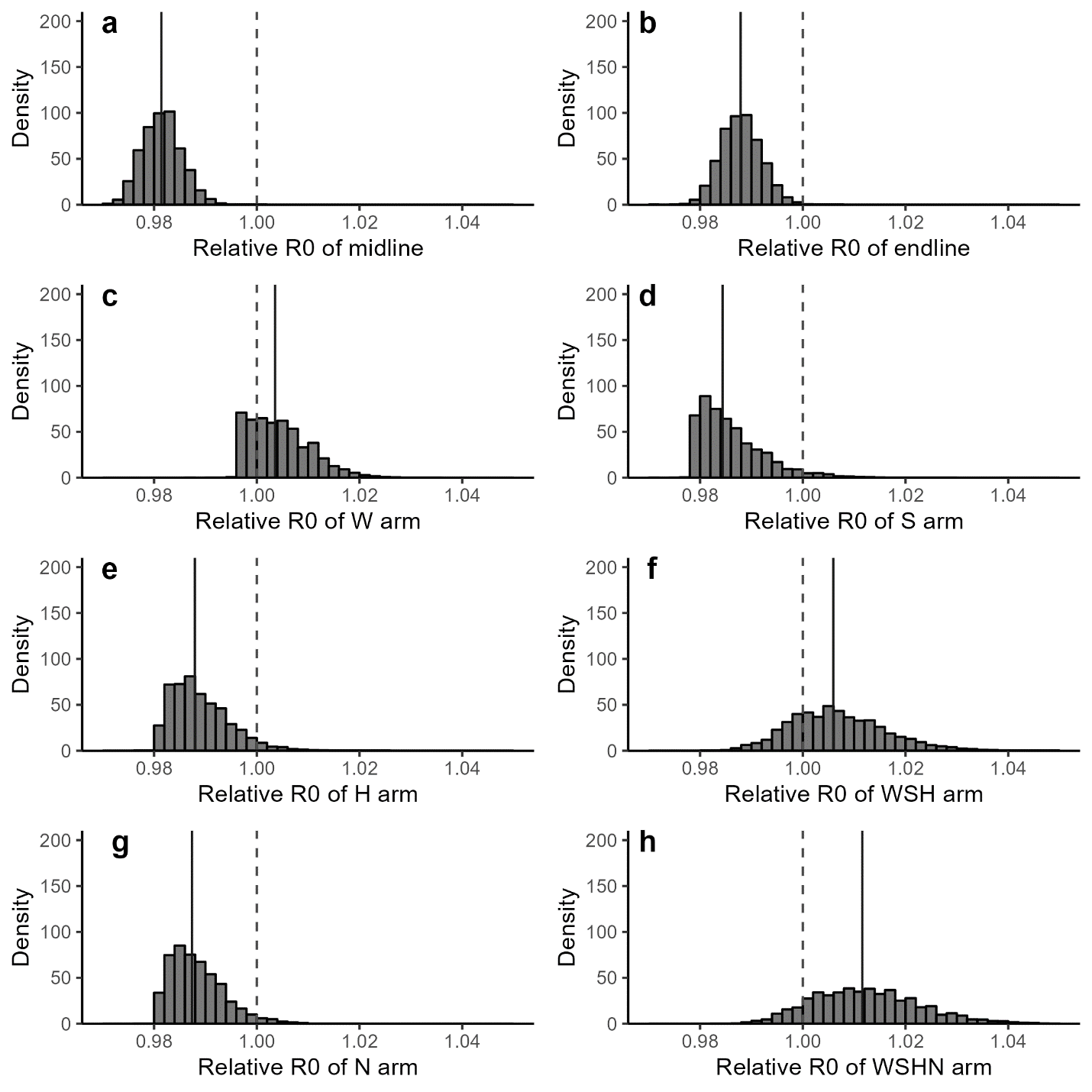

**Figure S3:** ***Time- and arm-specific relative basic reproduction numbers.*** *Distribution of timepoint- and arm-specific relative basic reproduction numbers (R0). The dotted line corresponds to 1.00, or no difference from the control arm at baseline. The solid lines give the mean values of the distributions.*

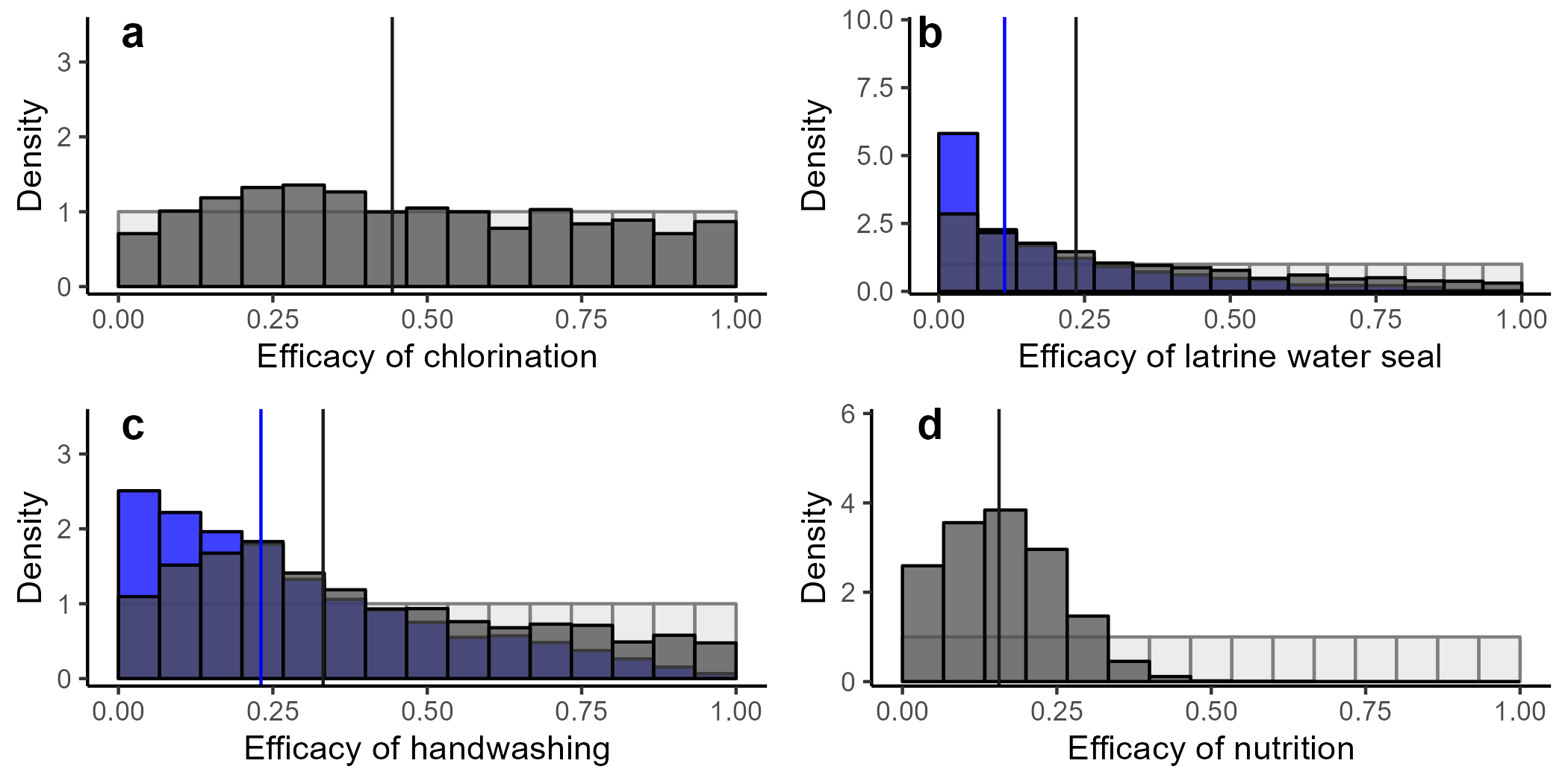

**Figure S4:** ***Efficacy****. The posterior distributions of the efficacy of (a) water chlorination, (b) latrine water seal, (c) handwashing, and (d) nutrition interventions are in dark gray. The posterior distributions of the efficacy of (b) latrine water seal and (c) handwashing preexisting conditions are in blue. Prior distributions (light gray) are given for all four sampled parameters. Vertical lines give median values of the intervention (dark gray) and preexisting conditions (blue).*

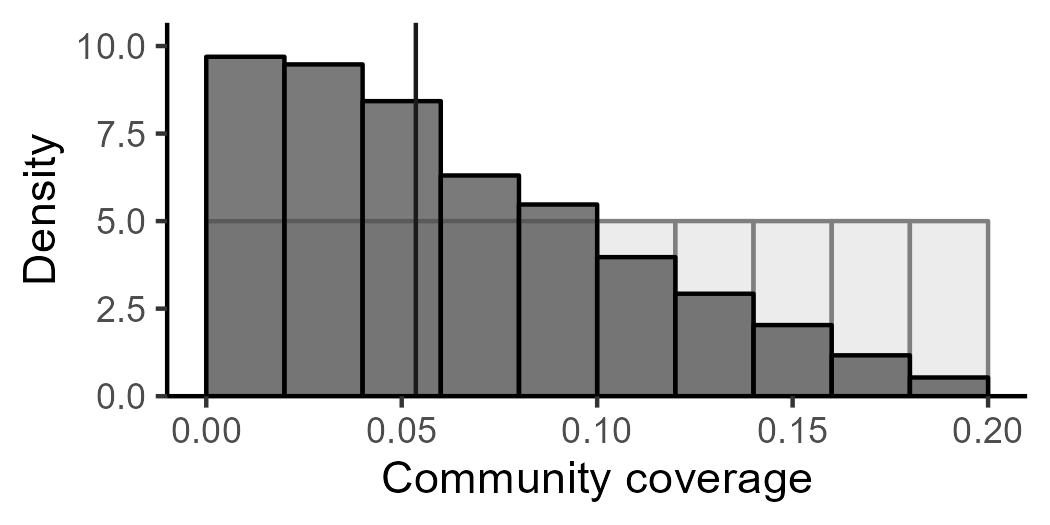

**Figure S4:** ***Coverage.*** *Posterior (gray) and prior (white) distributions for the estimated*

*fraction of the population enrolled in the study.*
